## Supplementary figures and images for "In depth analysis of patients with severe SARS-CoV-2 in sub-Saharan Africa demonstrates distinct clinical and immunological profiles"

### Supplemental figures

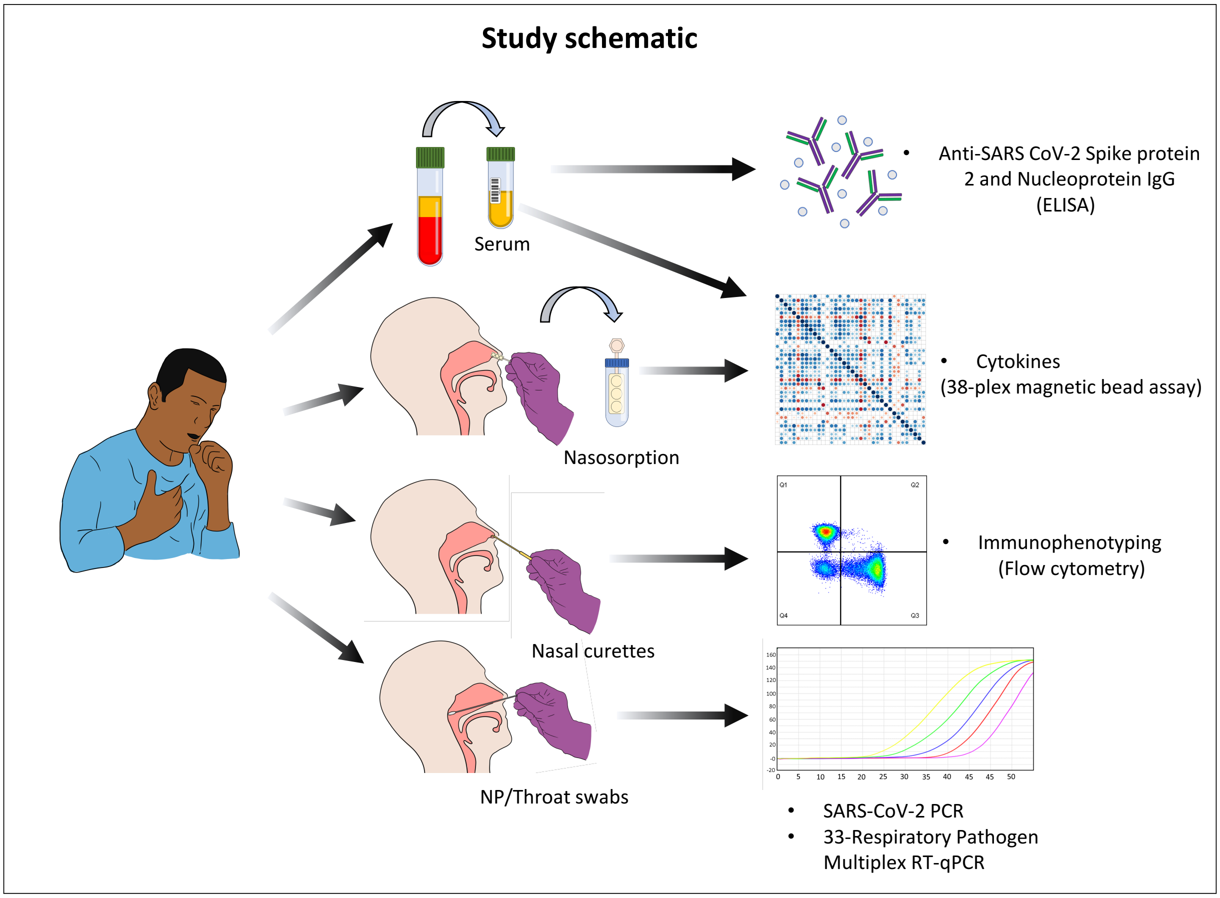

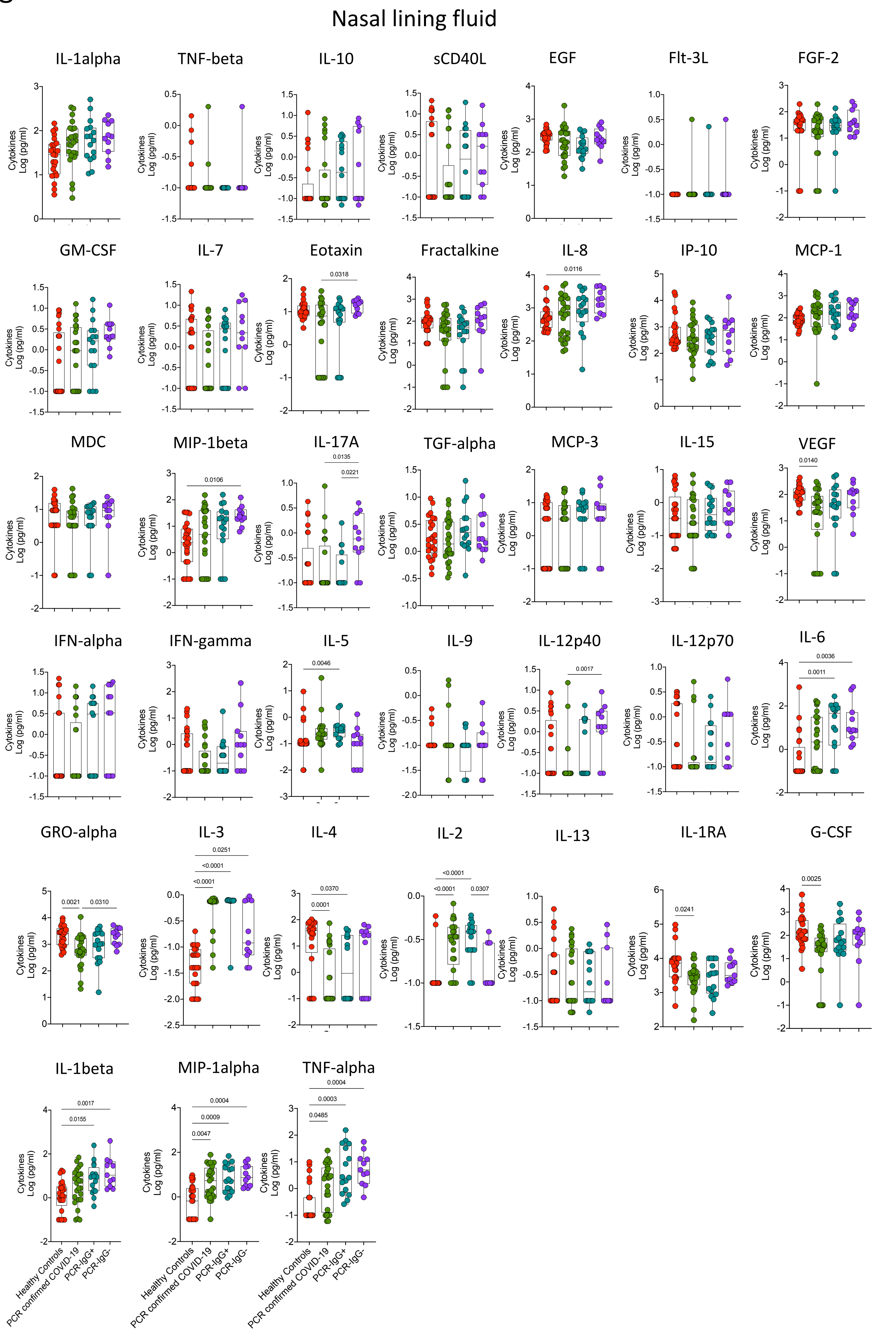

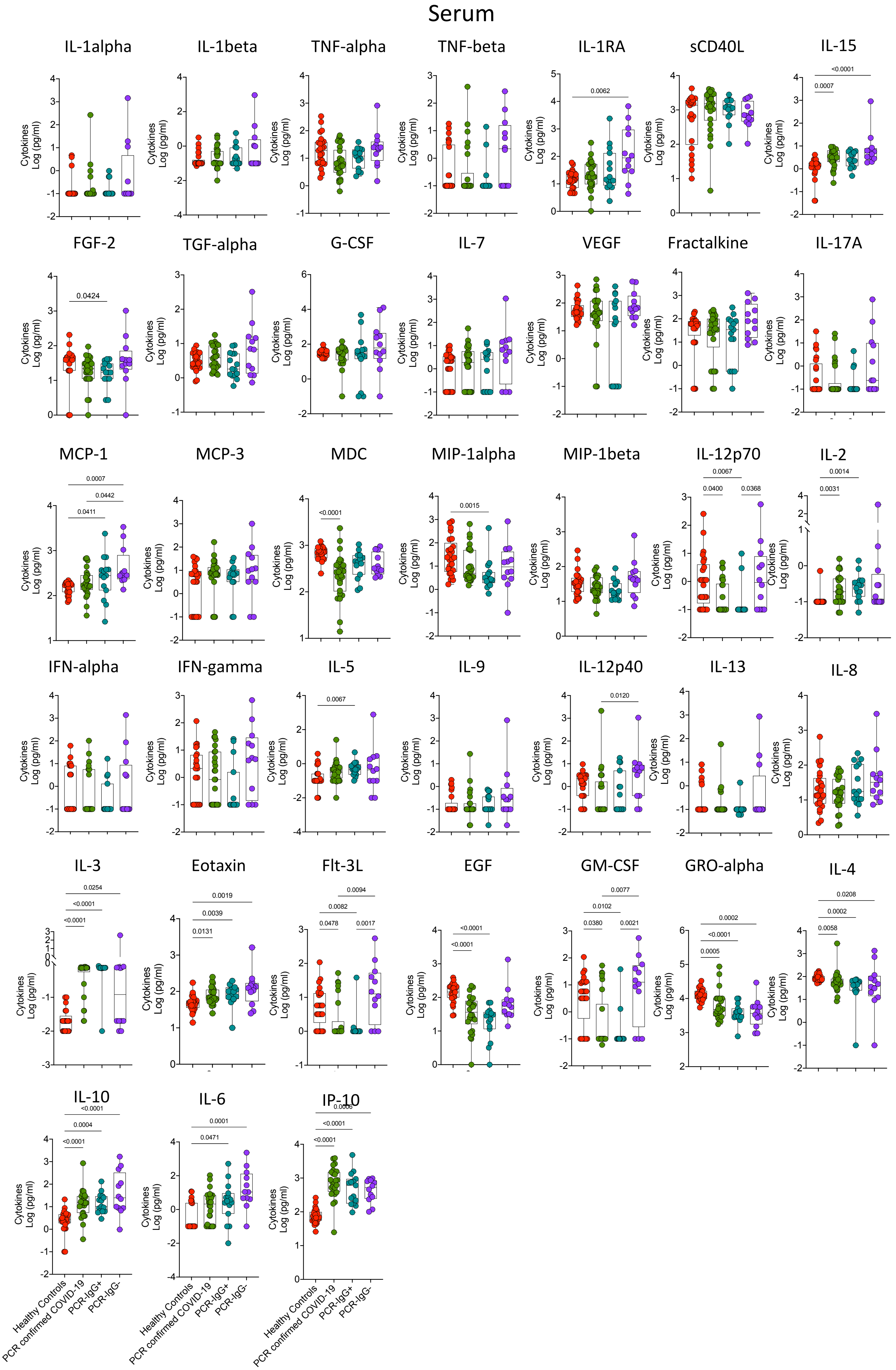

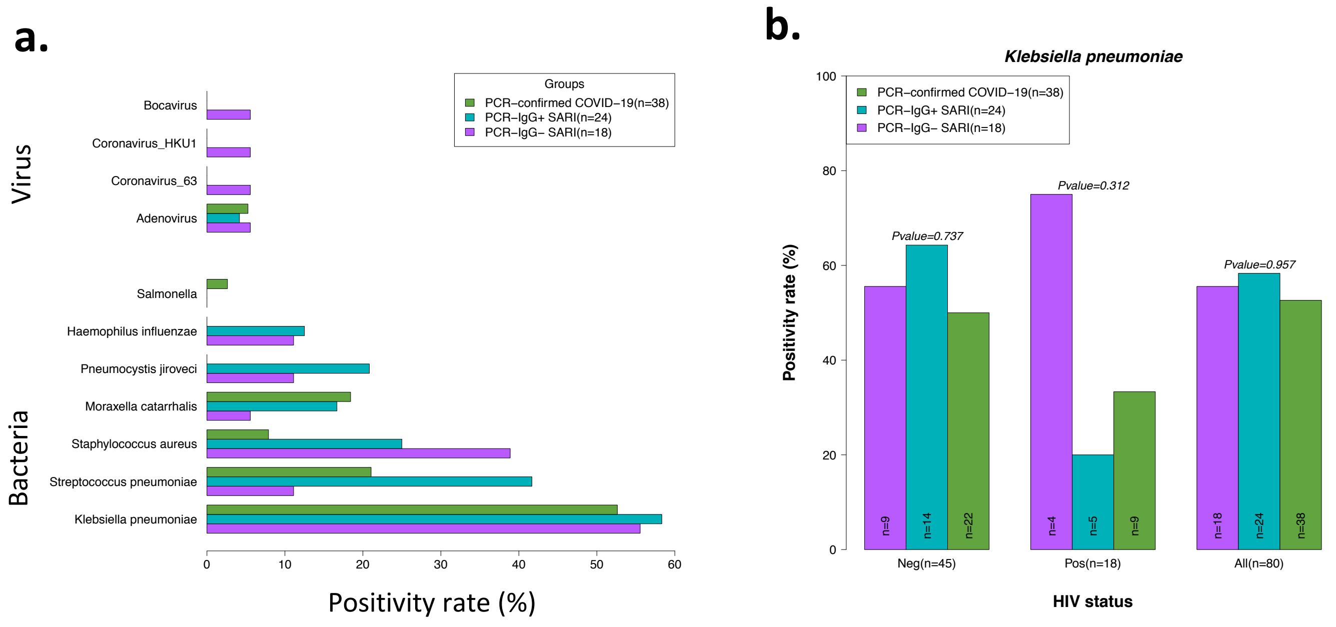
